# Ethnics and economics in COVID-19: Meta-regression of data from countries in the New York metropolitan area

**DOI:** 10.1101/2020.05.22.20110791

**Authors:** Hisato Takagi, Toshiki Kuno, Yujiro Yokoyama, Hiroki Ueyama, Takuya Matsushiro, Yosuke Hari, Tomo Ando

## Abstract

Ethnics and economics may affect prevalence and case fatality of Coronavirus disease 2019 (COVID-19). To determine whether COVID-19 prevalence and fatality are modulated by ethnics and economics, meta-regression of data from the countries in the New York metropolitan area were herein conducted. We selected 31 countries in the New York metropolitan area. 1) Prevalence and case-fatality rates of confirmed COVID-19 cases on May 20, 2020 and 2) income and poverty estimates were obtained in each country. We performed random-effects meta-regression using OpenMetaAnalys. The covariates included 1) black (%), 2) Hispanic or Latino (%), 3) poverty rates (%), and 4) median household income ($). Statistically significant (*P* < .05) covariates in the univariable model were together entered into the multivariable model. A slope (coefficient) of the univariable meta-regression line for COVID-19 prevalence was not significant for household income (P = .639), whereas the coefficient was significantly positive for black (coefficient, 0.021; *P* = .015), Hispanic/Latino (0.033; *P* < .001), and poverty (0.039; *P* = .02), which indicated that COVID-19 prevalence increased significantly as black, Hispanic/Latino, and poverty increased. The multivariable model revealed that the slope was significantly positive for only Hispanic/Latino (*P* < .001). The coefficient in the univariable model for COVID-19 fatality, however, was not significant for all the covariate. In conclusion, black, Hispanic/Latino, and poverty (not household income), especially Hispanic/Latino independently, may be associated with COVID-19 prevalence. There may be no association of black, Hispanic/Latino, poverty, and household income with COVID-19 fatality.

## Introduction

“New York City residents from low-income communities have tested positive for COVID-19 [Coronavirus disease 2019] antibodies at a higher-than-average rate, underscoring the disproportionate impact of the disease on people of color, Governor Andrew Cuomo said on Wednesday [May 20, 2020] (https://www.reuters.com/article/us-health-coronavirus-usa-new-york/new-york-citys-low-income-minority-areas-hit-hardest-by-covid-19-cuomo-says-idUSKBN22W2IG).” Ethnics and economics may affect prevalence and case fatality of COVID-19. To determine whether COVID-19 prevalence and fatality are modulated by ethnics and economics, meta-regression of data from the countries in the New York metropolitan area were herein conducted.

## Methods

We selected 31 countries in the New York metropolitan area, i.e. the New York-Newark, NY-NJ-CT-PA Combined Statistical Area (https://www.whitehouse.gov/wp-content/uploads/2020/03/Bulletin-20-01.pdf). 1) Prevalence and case-fatality rates of confirmed COVID-19 cases on May 20, 2020 from the Johns Hopkins Coronavirus Resource Center (https://coronavirus.jhu.edu/us-map) and 2) income and poverty estimates from the U.S. Census Bureau (https://www.census.gov/data-tools/demo/saipe/#/?map_geoSelector=aa_c) were obtained in each country (**Table**). We performed random-effects meta-regression using OpenMetaAnalyst (http://www.cebm.brown.edu/openmeta/index.html). A meta-regression graph depicted COVID-19 prevalence or fatality (plotted as logarithm transformed prevalence or fatality on the y-axis) as a function of a given covariate (plotted on the x-axis). The covariates included 1) black (%), 2) Hispanic or Latino (%), 3) poverty rates (%), and 4) median household income ($). Statistically significant (*P* < .05) covariates in the univariable model were together entered into the multivariable model.

## Results

Results of the meta-regression were summarized in **Supplementary Table**. A slope (coefficient) of the univariable meta-regression line for COVID-19 prevalence was not significant for household income (*P* = .639), whereas the coefficient was significantly positive for black (coefficient, 0.021; *P* = .015; **Figure, part A**), Hispanic/Latino (coefficient, 0.033; *P* < .001; **Figure, part B**), and poverty (coefficient, 0.039; *P* = .026; **Figure, part C**), which indicated that COVID-19 prevalence increased significantly as black, Hispanic/Latino, and poverty increased. The multivariable model revealed that the slope was significantly positive for only Hispanic/Latino (coefficient, 0.035; *P* < .001). The coefficient in the univariable model for COVID-19 fatality, however, was not significant for all the covariates (**Supplementary Table**).

## Discussion

The present meta-regression suggests that black, Hispanic/Latino, and poverty (not household income) may be positively associated with COVID-19 prevalence. Especially, Hispanic/Latino may be “independently” associated with COVID-19 prevalence. According to the most recent (19 May, 2020) data of the NYC Health, the age-adjusted case/death rate per 100,000 people was higher (approximately 1.5/2.0-fold for the case/death rate) in Hispanic/Latino (1307/217 of the case/death rate) and black/African-American (1470/209 of the case/death rate) than in White (934/104 of the case/death rate).^1^ The case/death rate per 100,000 people was also higher (approximately 1.5/2.3-fold for the case/death rate) in very-high poverty (2533/238 of the case/death rate) than in low poverty (1714/102 of the case/death rate).^1^ These findings may partially strengthen the present results. It is unclear 1) why Hispanic/Latino (neither black nor poverty) is independently associated with COVID-19 prevalence or 2) why none of black, Hispanic/Latino, and poverty is associated with COVID-19 fatality, which demonstrated in the present study. Disparity of racial and ethnic health should be identified and addressed in the pandemic of COVID-19.^2^ Furthermore, long-term legislation to improve social welfare would be introduced to address vulnerability in subjects with the most economical disadvantage.^3^

In conclusion, black, Hispanic/Latino, and poverty (not household income), especially Hispanic/Latino independently, may be associated with COVID-19 prevalence. There may be no association of black, Hispanic/Latino, poverty, and household income with COVID-19 fatality, which should be confirmed by further large hospital-based studies investigating whether these covariates are associated with COVID-19 fatality.

## Data Availability

The datasets generated during and/or analysed during the current study are available from the corresponding author on reasonable request.

## Conflict of Interest Disclosures

None reported.

## Figure legends

**Figure.**
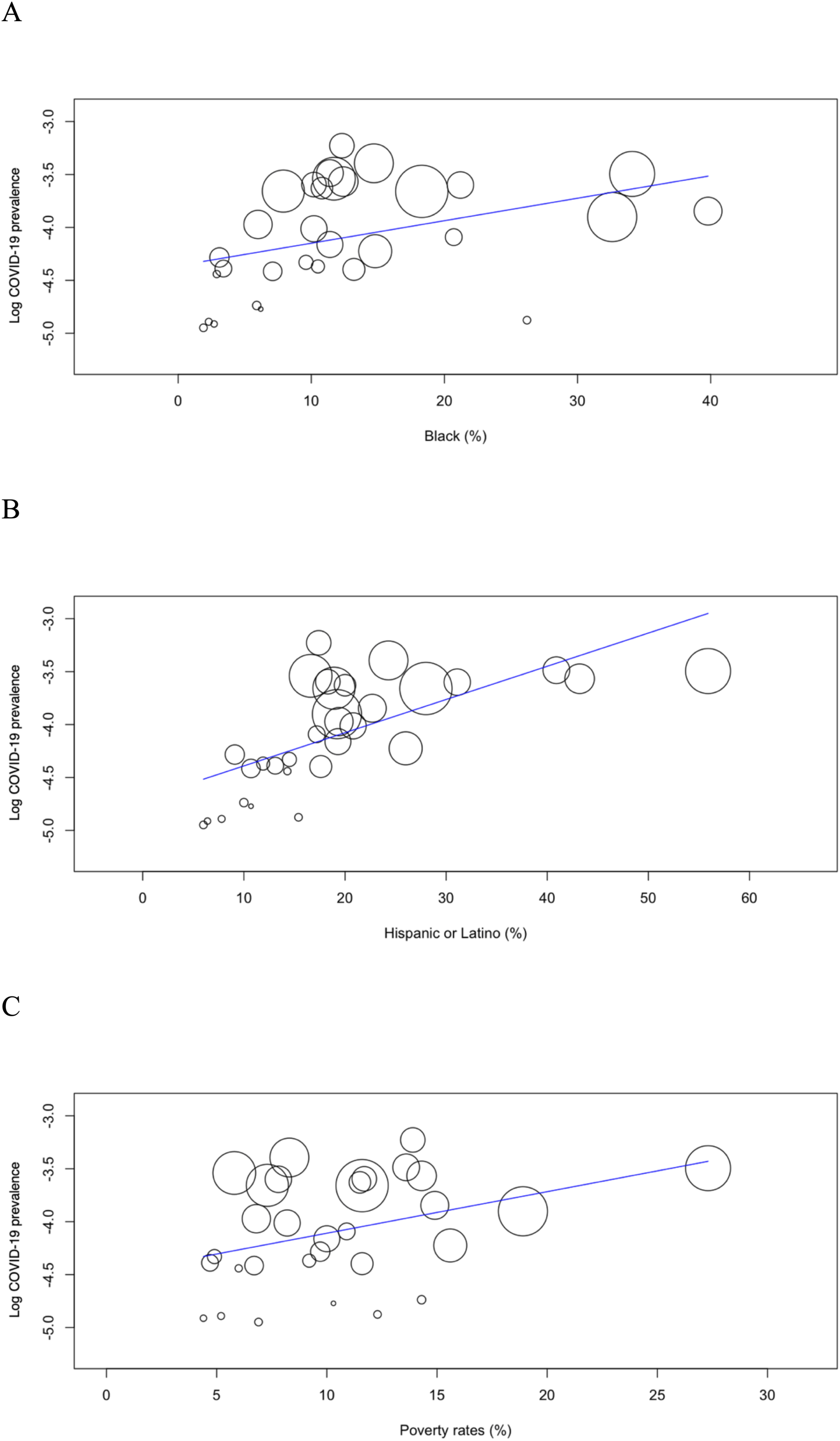
Meta-Regression Graph Depicting the COVID-19 Prevalence (Plotted as the Logarithm Transformed Prevalence on the Y-Axis) as a Function of a Given Factor (Plotted on the X-Axis)

**Table.**
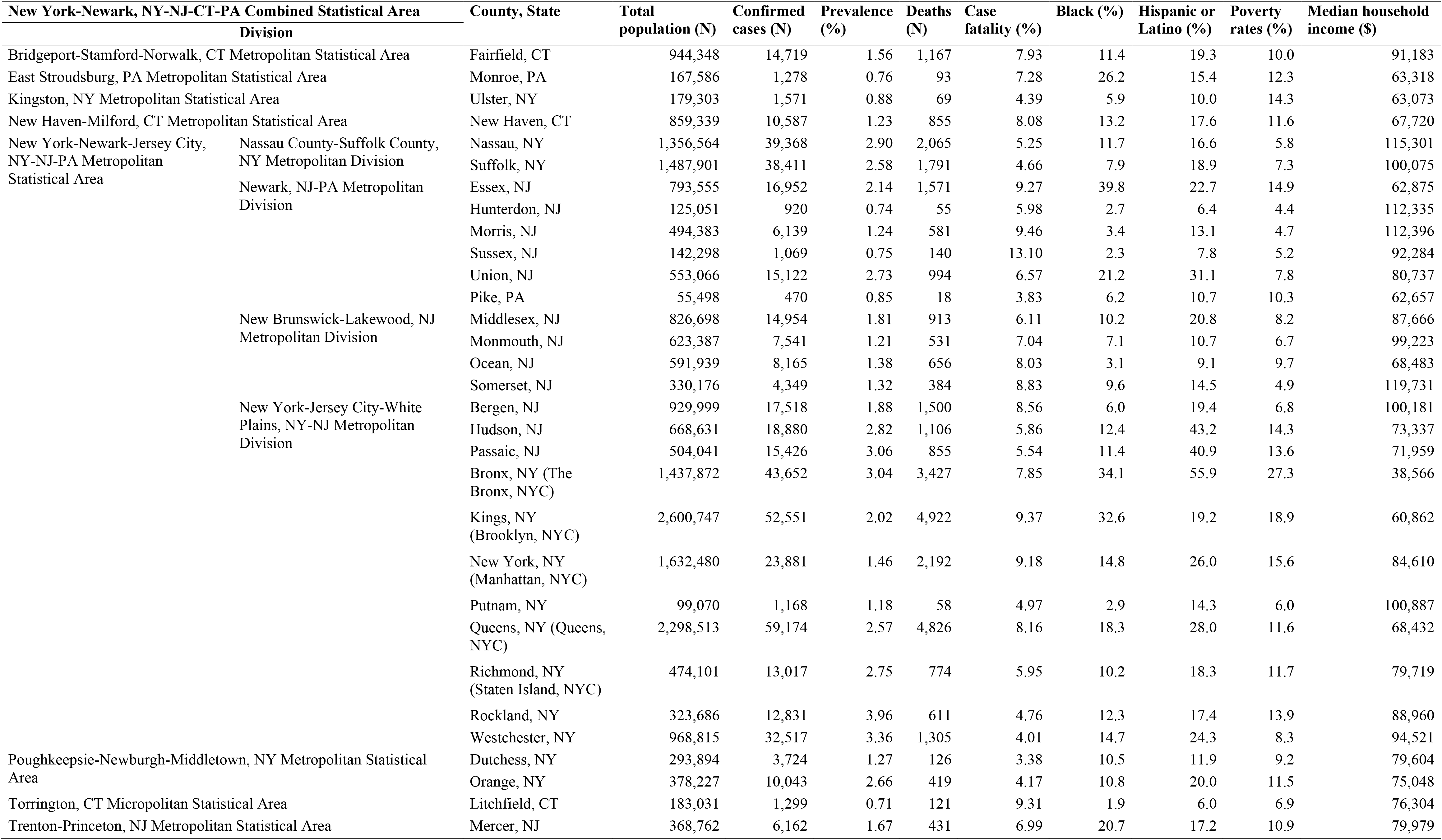
COVID-19 Prevalence/Fatality and Ethnics/Economics in Each Country in the New York Metropolitan Area (New York-Newark, NY-NJ-CT-PA Combined Statistical Area)

**Supplementary Table.**
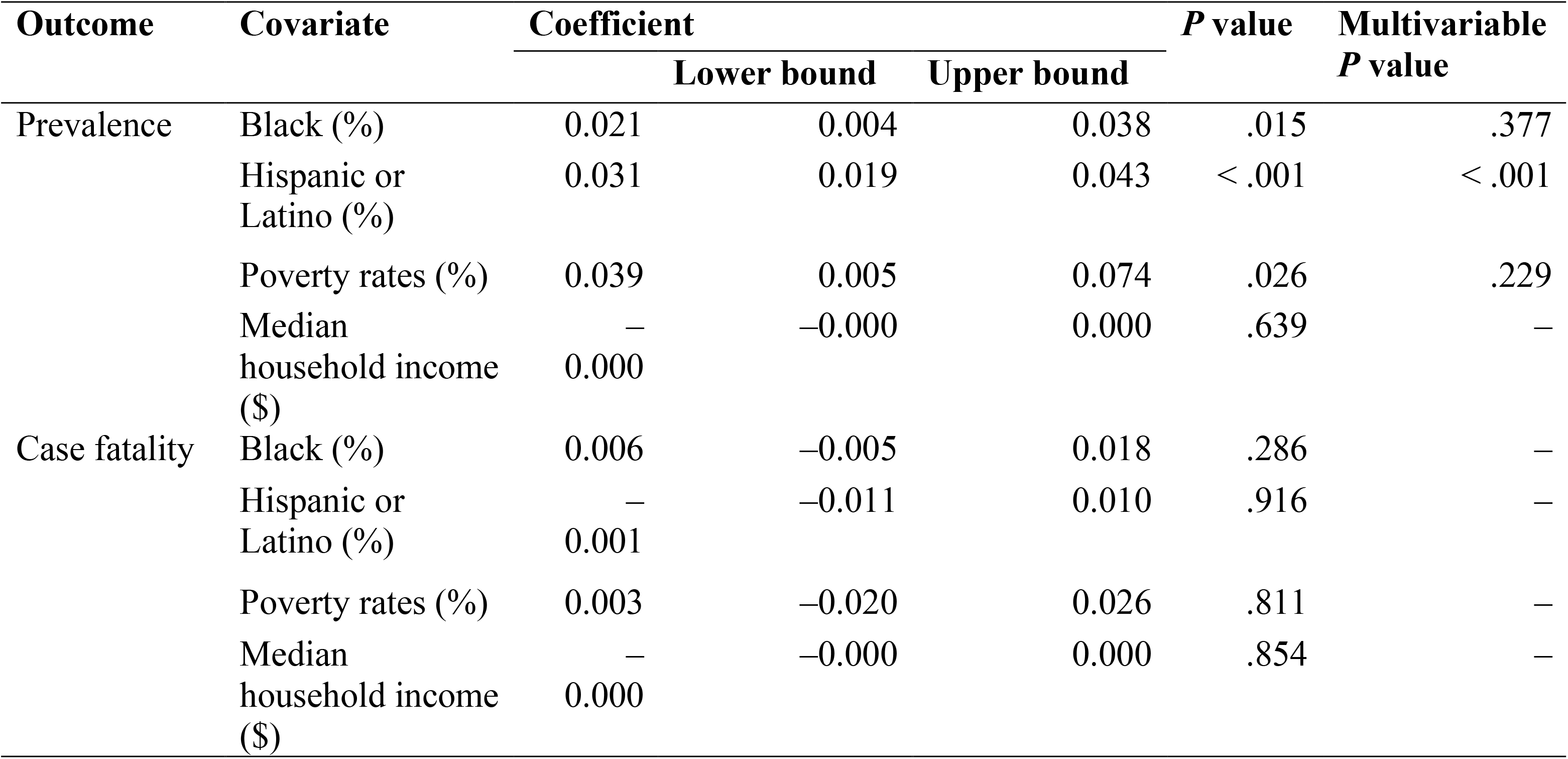
Summary of Meta-Regression

## Notes

### Competing Interest Statement

The authors have declared no competing interest.

### Funding Statement

No external funding was received.

### Author Declarations

No necessary IRB and/or ethics committee approvals have been obtained.

